# Adding Structure to Statistics: A Study on COVID-19 Dynamics in India

**DOI:** 10.1101/2020.05.26.20113522

**Authors:** Arundhati Dixit, Sarthak Vishnoi, Sourabh Bikas Paul

**Affiliations:** Indian Institute of Technology Delhi

**Keywords:** Time series forecasting, COVID-19, India, Regression Model, Holt Exponential Smoothing, Compartmental Model (SEIR)

## Abstract

This study pertains to COVID-19 in India, and begins by uncovering the statistical relationship between three time series-number of cases, number of deaths, and number of tests each day, using structural vector autoregression. Further, impulse responses of the before-mentioned series are studied. Effect of temperature and humidity on number of cases is analysed using the fixed effects model on city-level panel data. The next model utilises exponential smoothing for forecasting and conjecture for identifying peak specific to this data is presented. Lastly, multiple iterations of compartmental modelling, possible scenarios, and effect of underlying assumptions is analysed. The models are used to forecast number of cases (regression for short term and epidemiological for long term). In the end, policy implications of different modelling exercises and ways to check the implication for policy planning are discussed.

## I. INTRODUCTION

India recorded its first COVID-19 case on January 30, 2020, and since then the numbers have steadily increased. As on May 25, 2020, a total of 144,941 cases have been reported in the country. Table I depicts COVID-19 statistics for some countries. Table II lists the same for some Indian states. Given the huge variation in reported statistics, and numbers failing to corroborate to a certain estimate of disease dynamics like death rate, we ask ourselves an important question: Do statistical anomalies arise because of under-reporting, or erroneous estimates because of the contagious and hard to trace nature of disease cause these? Other possible reasons outside our scope for these observations might be diversity in disease mutation across geographies or difference in immunity and healthcare for individuals.

**Table I.**
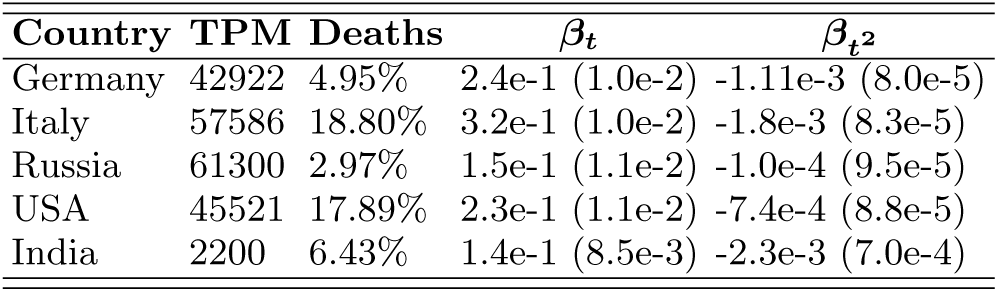
Worldwide statistics as on May 25, 2020 [4][7]

**Table II.**
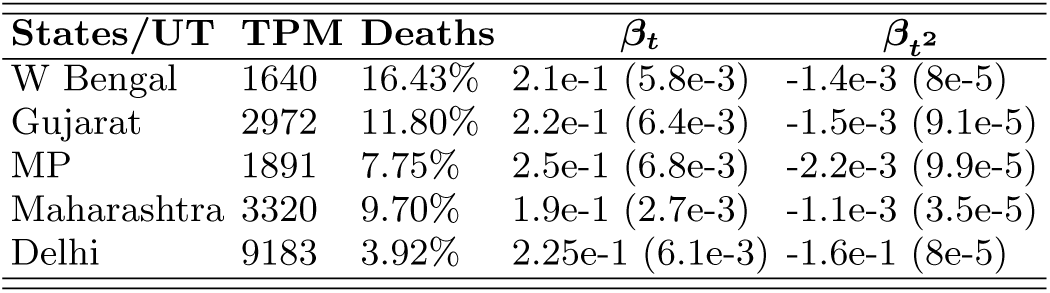
India, state-wise as on May 25, 2020 [5] Deaths are defined as % of number of deaths in closed cases, and TPM stands for Tests Per Million population. Parameter estimates are for log no. of cases regressed on time *t*, and its square *t*^2^, and brackets report standard errors

This paper is motivated by the need for understanding the relationship between time series of total number of positive cases, testing done and number of deaths in India in order to know how and when does one affect the other. This produces better forecasts, and aids in healthcare, policy planning and implementation, and would help in the study of such similar epidemic study if and when needed. We utilise the concept of structural vector autoregression (SVAR) to uncover interdependence, produce short term forecasts, and look at effects of other relevant factors like increased testing facilities, and temperature and humidity. In addition to this, we analyse the usefulness of exponential smoothing model for short term forecasts, and propose observational conjectures based on the same. Using relative trend of actual and forecast numbers, we propose a method for identification of having reached the peak number of cases. We strengthen short term forecasts by defining bounds using a committee of models. Lastly, we utilise iterations of long term epidemiological model, SEIR to comment on lockdown policies and its effect on long term disease dynamics. We propose a modification in the classical model which captures the current scenario.

This study tests several conjectures and adds to the existing literature on statistical analysis of COVID-19 cases. It helps produce richer predictions of number of cases in India. The study also proposes suitable modifications in using econometric methods to understand evolution of disease in the country. More details about the modelling can be found in the **online appendix**.

## II. RELATED WORK

A number of statistical studies for COVID-19 cases have been carried out and would be underway. Regression, exponential smoothing and compartmental modelling are standard procedures for forecasting.

The study by Deb and Majumdar [2020] analyzed the number of cases per million population in various provinces of China using an ARIMA model with a structural break. Qi et al. [2020] showed the effects of Temperature and Relative Humidity on the number of confirmed cases in cities across SE Asia and China. Gupta et al. [2020] have studied the trends for the underlying time series for the number of Confirmed Cases in India for the early part of the spread (till March) in the country. The study published by Petropoulos and Makridakis [2020] utilises exponential smoothing to estimate forecasts for cumulative daily figures aggregated globally of three variables: confirmed cases, deaths and recoveries. The study by Yang et al. [2020] has used SEIR modelling with assumptions and scenario tailored to the situation in China to forecast disease dynamics. Prem et al. [2017] published a study, capturing heterogeneities in contact networks, and the effect of the same in epidemic models. Specific to India, Singh and Adhikari [2020] have studied modelling lockdown scenarios based on SIR modelling.

## III. DATA AND IMPLEMENTATION

All the data [5] [8] compiled and condensed by us can be found on this **repository**. The link also has the coes that have been used to generate results presented in the submission. All the methods are implemented using RStudio version 1.2.5033, along with R version 3.5.1. The data used is till May 20, 2020.

## IV. REGRESSION ANALYSIS

### A. How are the three time series inter related?

For India on a particular day, we consider *x_t_*- the number of confirmed COVID-19 cases; *y_t_*- number of deaths; *z_t_*- number of samples tested; *t*- time. Figure 1 shows the evolution of the three time series.

**FIG. 1.**
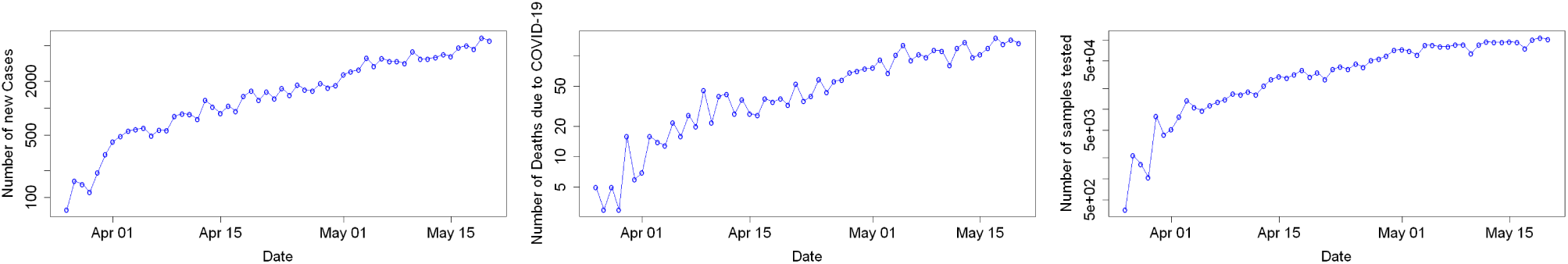
The three time series, a) *x_t_*, *b*) *y_t_*, *c*) *z_t_*

Intuitively, we may expect some correlation between these three time series, and use the same to define our model. For instance, an increase in confirmed cases should lead to an increase in testing in the country. Increase in testing potentially leads to an increase in COVID-19 cases contemporaneously. Increase in number of deaths due to the disease should make the authorities more proactive, and hence more testing will be done to identify the patients in the coming days. Increase in number of confirmed cases should also lead to an increase in number of deaths in the country due to COVID-19. In order to check these conjectures, we construct the model for the three time series using structural vector autoregression (SVAR). We test for stationarity of series, and conclude that all three series are trend stationary, hence during our formulation we also include time as a regressor. Models with different degrees of time variable on the basis of AIC values are compared in the paper by Deb and Majumdar, and *t* and *t*^2^ are found to be best suited, hence used here. Based on the PACF plots and AIC values, we include lagged values (lag = 3) for the regressand. We regress on log transformation of variables.

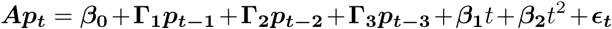

Here, ***A***, **Γ**_1_, **Γ**_2_ and **Γ**_3_ are *M*_3×3_ matrices. ***p_t_*** is *(log(x_t_*), *log(y_t_*), *log(z_t_*))*^T^* and *β****_i_***s are the coefficients for time trends and the constant term.

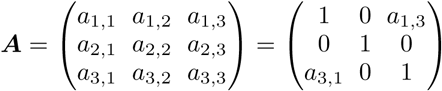

***A*** determines the structure of contemporaneous relationship between the three time series. We apply some restrictions to the elements of ***A*** to compute it deterministically. The diagonal entries in ***A*** are 1 as each variable affects itself. *a*_1,2_ determines the effect of deaths on the number of confirmed cases contemporaneously and *a*_2,1_ the opposite of this. These two elements should be zero because there is no reason as to why one should affect the other at the same time. The same holds for *a*_2,3_ and *a*_3,2_ which are the contemporaneous relationship between the deaths and the samples tested. As discussed above, more testing leads to higher number of confirmed cases, hence, *a*_1,3_ and *a*_3,1_ cannot be zero. So we have two elements of the ***A*** which need to be estimated.^1^

## Results

Figure 2 shows the Forecast Error Variance Decomposition of the model described above. We see that a large amount of variance in errors of confirmed COVID- 19 cases (*x_t_*) comes from the number of deaths (*y_t_*) and the number of samples tested (*z_t_*), while the same for *z_t_* comes from *x_t_* and *y_t_*. We observe a different scenario for *y_t_*, where the variance in errors solely depends on itself. Thus, from Figure 2 we can describe a weak causal relationship for our three time series, where *x_t_* and *y_t_* cause *z_t_*, *y_t_* and *z_t_* cause *x_t_* and *y_t_* causes itself. We establish that SVAR modelling of the three time series is significant. Please refer to the **online appendix** for the estimates of various parameters.

**FIG. 2.**
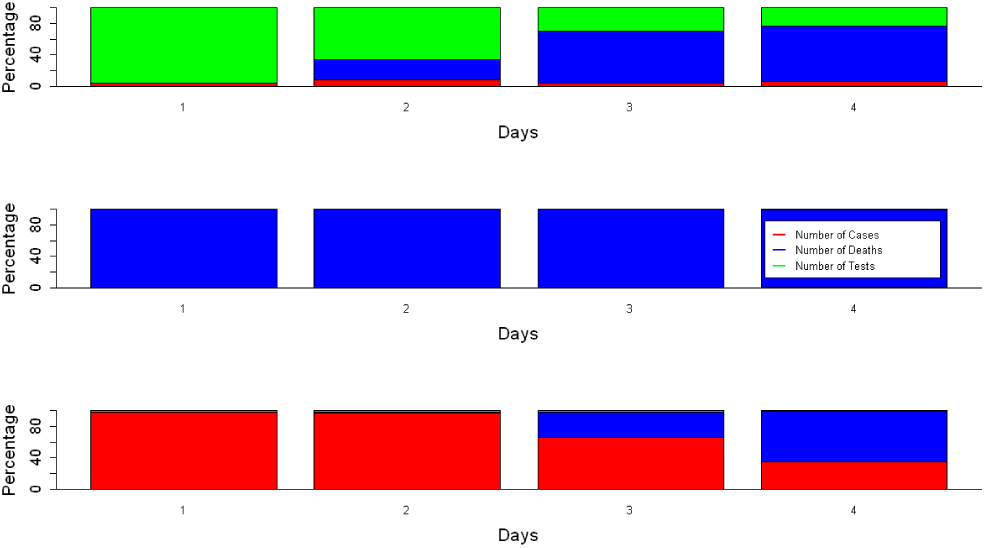
Forecast Error Variance Decomposition of the three time series

## B. What do we know about the Impulse Responses?

One way to interpret the results of SVAR is to see the impulse response functions. In this section we talk about the responses in number of confirmed cases when an impulse is injected to the testing numbers. The other impulse responses can be found in the **online appendix**. News articles [2][1] report that more than 75% of the confirmed cases are asymptomatic which can only be detected by testing. Table I shows that the testing levels in India are very low compared to the Western countries, and even though we see a gradual increase in *z_t_* over time in Figure 1, this is not enough. This raises concerns over the actual number of COVID-19 cases in the country which might be higher than the confirmed cases reported by relevant agencies.

We discuss the impact of doubling the testing capacity for two cases. In the first case, we do it for a single day, while in the second case, we maintain the increased testing for a prolonged period. The first case results in an increase of 9.8% in confirmed cases for COVID-19 contemporaneously which goes back to zero in about a week’s time. The second case leads to a steady rise of 13.7% in confirmed COVID-19 cases for a week and then becomes constant at that level. This can be seen in Figure 3. Increased testing is evidently is beneficial, as we are then able to identify and treat more cases earlier. This also helps in curbing the spread of the disease as accruing cases are isolated from the general population, thus bringing down the *R*_0_ value, the details of which are in Section VII.

**FIG. 3.**
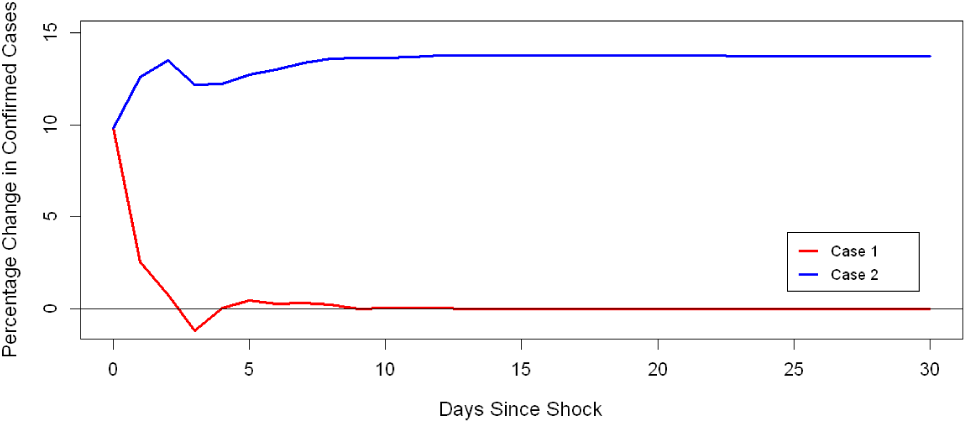
Impulse response functions for the two cases described

## V. METEOROLOGICAL FACTORS

### Do Temperature and Humidity affect the number of COVID-19 cases?

There is variation in factors like temperature and humidity over geographies and also over time since January 30, which was when the first case was detected. These meteorological factors are exogenous with their own trend in nature. We want to look at the effects that this may or may not have on the number of cases in India, and so we use fixed effects model, thus taking into account heterogeneity at city level and getting the relationship with temperature, humidity and time nationally.

Studies in different places (Bashir et al. [2020] in New York, Qi et al. [2020] in China and §ahin [2020] in Turkey) have found significant relationships between the number of COVID-19 cases and meteorological factors. We conduct a study for five cities-Indore, Ahmedabad, Delhi, Jaipur and Mumbai.^2^ These have different geographical features, climate and high number of confirmed COVID-19 cases. We check for effect of Temperature and Relative Humidity on the same. We observe some degree of correlation, and lag of 3 for moving average values of temperature and humidity (window size of 4 (including contemporaneous point and 3 previous days’ values)) is taken for this analysis. In addition, a linear and quadratic time trend is included.

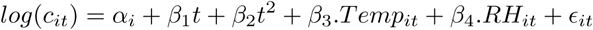

In the above equation, *i* is for a city and *α_i_* is the fixed effect of that city. *c_it_* is the number of confirmed COVID- 19 cases, *Temp_it_* is moving average of temperature over four days, and *RH_it_* is moving average of Relative Humidity over four days. The significance of coefficients is tabulated in Table III. The fixed effects of each city can be found in **online appendix**

**Table III.**
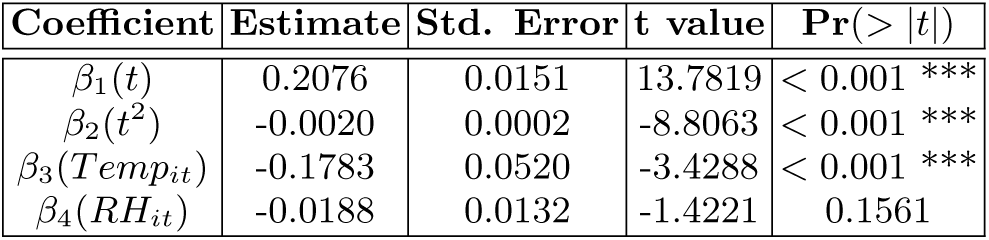
Fixed Effects Model estimates for log(*c_it_*)

### Results

Table III shows a significant coefficient for moving average of temperature, which is negative. This is in coherence with the other studies which had found temperature to be negatively significant in different places. As we have controlled for time, we conjecture that the expected number of COVID-19 cases in a city with higher temperature will be lower than the expected number of cases in another city with a lower temperature on the same day, provided the fixed effects of these two cities are insignificantly different. We fail to find any significant relationship with Relative Humidity.

## VI. EXPONENTIAL SMOOTHING

Similar to regression, exponential smoothing is a live forecasting technique. In regression, we permit for a finite lag, whereas here, all weighted past data points are utilised. There is a clear time trend in data albeit no seasonality, so Holt smoothing is implemented.

Forecast equation: 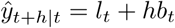

Level equation: *l_t_* = *αy_t_* + (1 − *α*)(*l_t_*_−1_ + *b_t−_*_1_)

Trend equation: *b_t_* = *β(l_t_ − l_t−1_*) + (1 − *β)b_t_*_−1_

Here, *l_t_* is the estimate of level of series at time *t*, *b_t_* is an estimate of trend (slope) of series at time *t*, *α* is the smoothing parameter of the level, 0 *< α <* 1, *β* is the smoothing parameter of trend, 0 *< β <* 1.

Forecasting exercise is performed, where the results are updated every 10 days beginning from March 12, and forecasts and actual numbers are plotted in Figure 4.

**FIG. 4.**
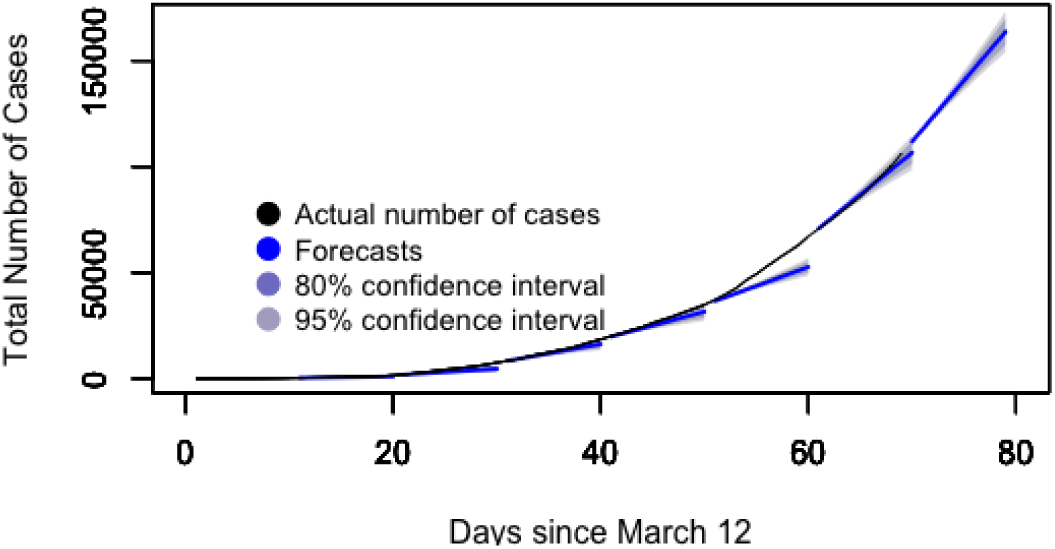
Forecasting exercise

### Observational proposition

The forecast values here give a lower bound on cases, and actual numbers top the predicted trend based on past values in general. So when the actual numbers begin meeting the forecast or are less than the latter, we can say that the prior trend has exhausted and as a result, we are either at the peak of number of cases, or the downturn in number of cases is expected in days to come.

Similar proposition holds for death statistics, the details of which can be found on the **online appendix**.

## VII. COMPARTMENTAL MODELLING

### A. How does length of lockdown change disease dynamics?

#### Classical SEIR

We employ a deterministic Susceptible Exposed Infectious Recovered (SEIR) model based on progression of disease, epidemiological status of individuals and intervention measures of disease transmission. Figure 6 depicts the model structure, and differential equations below are solved to get solutions for certain cases that are plotted in Figure 5.

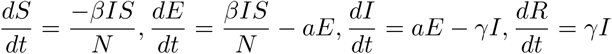

*S* + *E* + *I* + *R* = *N*, *N* is the total population.

**FIG. 6.**
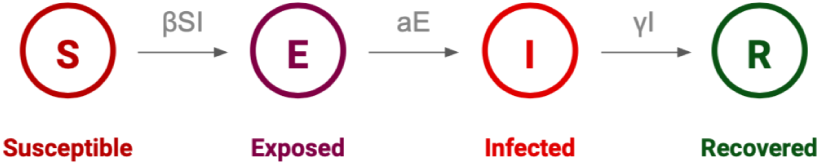
Population compartments & model parameters

**FIG. 5.**
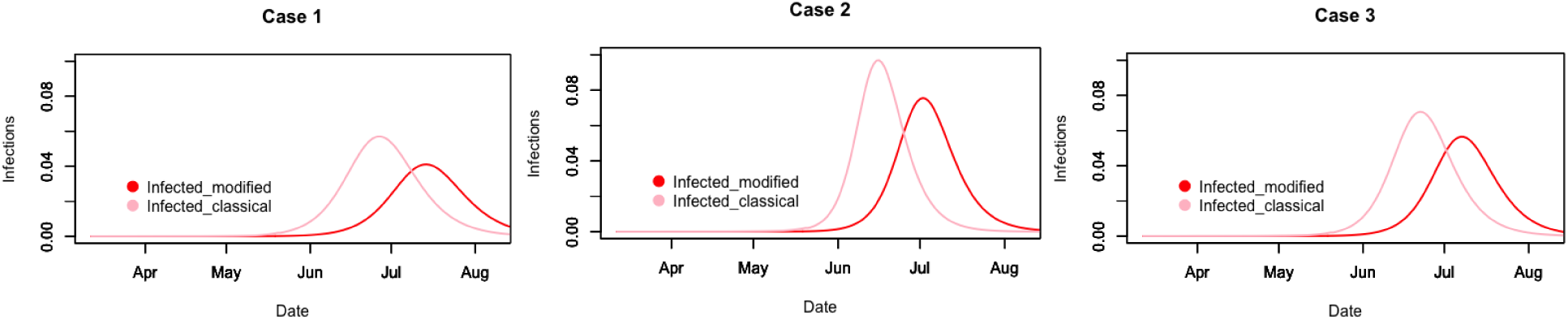
Plots of case-wise analysis of SEIR modelling, classical and modified, y-axis is as a fraction of the whole population

**Assumptions:**

1. Total population is constant, natural birth and death rates neglected, no new individuals travel in.
2. Latent and infectious periods of the virus are exponentially distributed.
3. Infectious individuals based on probabilistic process either recover or die and become removed class, and no more participate in the SEIR chain.
4. Population is homogeneously well mixed.
5. Every individual belongs to one of the classes.

An important concept here is that of **reproduction number**, 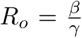. This number captures the dynamics of disease spread in the population. A strong assumption is for it to be a constant for the country, as it would differ because of varying geographies and demographics, but that is how we have done it in this study. Estimates of *R_o_* are a potential way of looking at impact and extent of any policy implementation like lockdown.

Constant parameters in the model [13]:

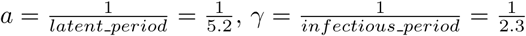

Three cases have been considered for plots:

*<u>Case I</u>*: The partial lockdown, as it is right now, is extended indefinitely (with an *R_o_* value of 2.2). Initial value of *R_o_* is taken to be 4, and lockdown period *R_o_* is 1.45, based on fitting actual data points for different periods of time for India, and then corroborating the estimate with other studies mentioned in Section II.

*<u>Case II</u>*: Partial lockdown ends by end of June, and everything reopens to near normalcy.

*<u>Case III</u>*: Partial lockdown opens to a more conservative normalcy post June (work from home and online classes wherever possible, restricted travel services).

68.84% population is rural and 31.16% is urban [6]. 28.6% of rural and 23.6% of the urban population is below 14 years of age. As on May 24, Maharashtra, Tamil Nadu, Gujarat and Delhi account for 67% of the total number of cases reported in country. Goa, Dadra & Nagar Haveli, Arunachal Pradesh, Mizoram, Lakshadweep, Nagaland, and Sikkim had one/very few cases that recovered, so we remove these states and UTs from our analysis, which make up 0.52% of the population. Also, we consider urban and 30% of rural population in the analysis.

### B. How do we account for population heterogeneity?

***Age structured SEIR:*** The structuring of model can be done in several ways by including age based or geo spatial data, or a combination of multiple, thus easing assumption (4). We have picked and demonstrated age structured compartmental model in this paper. Figure 7 presents the altered model, where we partition the compartments and solve the listed differential equations. The two partitions *i* and *j* are taken to be senior citizens (≥ 60 years age, comprising 8.5% of the total population), and everyone else respectively. We have demonstrated one case in Figure 8, where the lockdown with relaxations ends by end of June, and restrictive normalcy follows.

**FIG. 7.**
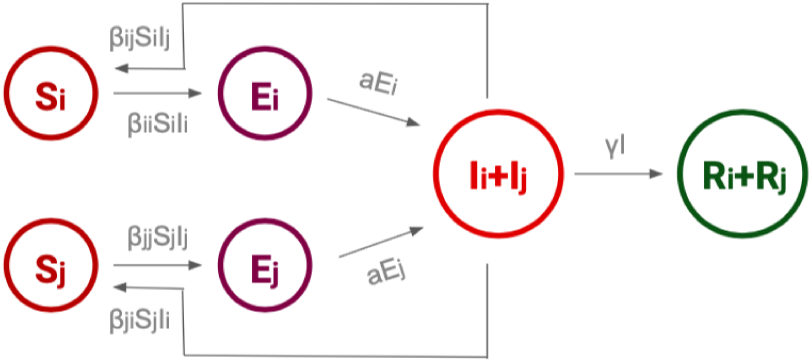
SEIR accommodating two age groups *j* and *i*

**FIG. 8.**
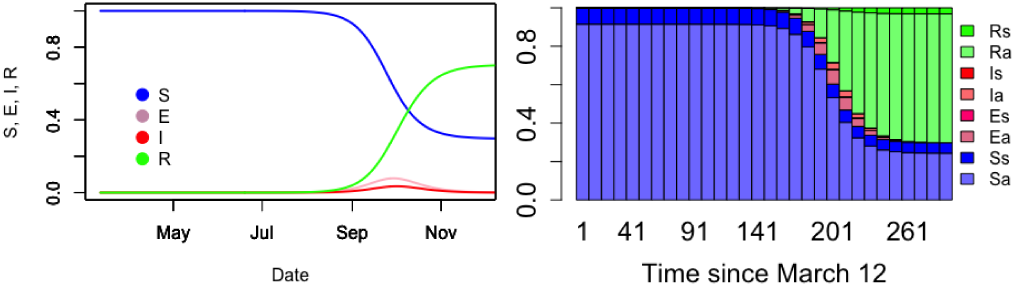
Results of SEIR model, a) time based chart, b) chart showing diseased population for both sections, *i* and *i*

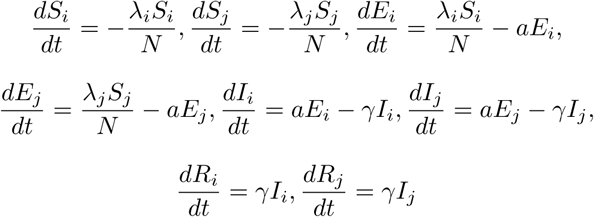

*λ* denotes the age specific force of infection, and

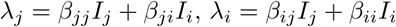

We see that while cases continue to rise, unplanned normalcy can lead to an increase in number of cases, thus rendering the lockdown futile. But we may expect effective contact tracing and quarantining of cases as we are better prepared to deal with the problem now.

### C. Can we do better than classical assumptions?

Here we ease assumption (5).

Proposition:

For a disease, the parameters *γ, a, λ* are a constant. But since the declared pandemic has caused countries to impose lockdowns, the contact pattern and hence *β* alters. The effect of lockdown is reflected in not just changing *β* in the model, but also in changing total level of susceptible population (which is S). This is also the policy implication-lockdown is meant to decrease contact and total population susceptible to the disease.

Continuing from classical model assumptions, we consider 10% of the rural population and urban aged above 14 during lockdown. For partial lockdown, we consider 80% urban and 20% rural aged above 14. For restrictive normalcy, we consider 85% urban and 20% rural population. Based on this model, we present case wise analysis in Figure 5 alongside classical SEIR.

**FIG. 9.**
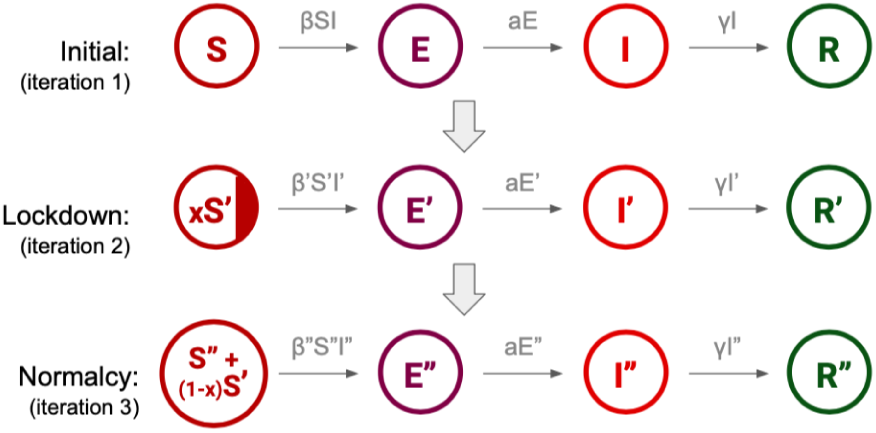
Evolving compartment SEIR

### D. Takeaways from SEIR

1. There is a trade off between peak value and time when peak arrives, and in order to contain that, social distancing, efficient contact tracing and isolation of cases is the key.
2. By plotting Figure 5, we study the case wise scenario, but we also look at the effect that assumptions has on the model as a whole. The results are always sensitive to the model assumptions, so we need to be careful before we accept or discard the forecasts of any model.
3. The curve from modified SEIR is flatter and delayed because by reducing susceptible population in multiple iterations, we have essentially bettered the societal response.
4. The total susceptible population is not zero, nor are the recoveries equal to 1. So although it is hard to comment on what we can anticipate, but even after the peak and downturn, virus does not die out.
5. Since actual data has not been worked with in this modelling, model fit or forecast sensitivity is not applicable in the conventional sense.

## VIII. FORECASTS AND ACTUAL VALUES

We present 5-day forecasts for the number of confirmed COVID-19 cases from different models in this paper. While regression provides an upper bound on number of cases, exponential smoothing gives linear output (here, 5757) which is a lower bound. Together they provide an interval of expected number of cases.

**Table IV.** 5-day forecasts from models

| Date | VAR | SEIR | Actual |
| --- | --- | --- | --- |
| May 21 | 6204 | 6164 | 6023 |
| May 22 | 6208 | 6628 | 6536 |
| May 23 | 6879 | 6831 | 6665 |
| May 24 | 7064 | 7210 | 7111 |
| May 25 | 7346 | 7532 | 6414 |

## IX. CONCLUSION

We construct a number of models and we see that the results from mathematical modelling are highly sensitive and specific to the choice of model and its underlying assumptions. We can do better with more data about demographic contact rates and lockdown effects, which we have demonstrated with ballpark figures in our paper. The forecasting exercises need to be updated periodically with data and substantiated hypotheses. Since our study is based on reported statistics, it does not account for virus properties and heterogeneity in time series because of its specific and geo spatial varying properties. Short term models help in efficient planning for testing and healthcare facilities, while long term predictions are the key to implementation of policies like lockdown and making resources available to the general population, a large proportion of which cannot work from home and depends on daily to monthly earnings. By studying both, we have tried to capture the entire picture of the pandemic. Asymptomatic cases are worrying as they can only be identified by contact tracing and testing, and without any symptoms, they are disease carriers. Use of technology to tackle problems will go a long way in improving conditions. Additionally, it is important in such a scenario for the unaffected population too to take precautionary steps to ensure that they are protected from any possible form of disease contraction. Meanwhile, the nation must brace itself for a long and hard battle, for every case and death is more than just a statistic and it is crucial to come together to prevent losses detrimental to the nation.

## Data Availability

All the data used is available in our repository, also included in the document itself.

https://github.com/SarthakVishnoi01/TSF-Challenge

## X. CONFLICTS OF INTEREST

We declare that we have no conflicts of interest.

## Footnotes

1 The sensitivity analysis for the off-diagonal elements of the A matrix showed that *a*_1,3_ and *a*_3;1_ are not sensitive to other values while the other elements were highly sensitive to the other estimated parameters

2 Indore: *SD_Temp_* = 2.051 (Standard Deviation of Temperature), *SD_RH_* = 8.261 (Standard Deviation of Relative Humidity) Ahmedabad: *SD_Temp_* = 1.819, *SD_RH_* = 8.283 Delhi: *SD_Temp_* = 3.737, *SD_RH_* = 8.164 Jaipur: *SD_Temp_* = 3.478, *SD_RH_* = 9.316 Mumbai: *SD_Temp_* = 1.497, *SD_RH_* = 6.107

## Notes

### Competing Interest Statement

The authors have declared no competing interest.

### Funding Statement

No external funding was received

### Author Declarations

All data is from publicly available domain

